# Insecticide Resistance Surveillance of Malaria and Arbovirus Vectors in Papua New Guinea 2017-2022

**DOI:** 10.1101/2022.05.01.22274242

**Authors:** Michelle Katusele, Solomon Lagur, Nancy Endersby-Harshman, Samuel Demok, Joelyn Goi, Naomi Vincent, Muker Sakur, Absalom Dau, Lemen Kilepak, Stephen Gideon, Christine Pombreaw, Leo Makita, Ary Hoffmann, Leanne J Robinson, Moses Laman, Stephan Karl

## Abstract

**Background:** Insecticide resistance monitoring is key for evidence-based control of *Anopheles* and *Aedes* disease vectors in particular, since the vast majority of insecticide-based public health adult vector control tools are reliant on pyrethroids. While widespread pyrethroid resistance in *Anopheles* species and *Aedes aegypti* has been described in many countries, data for Papua New Guinea are scarce. Available data indicate the local *Anopheles* populations remain pyrethroid-susceptible, making regular insecticide resistance monitoring even more important. Knowledge on *Aedes* insecticide resistance in PNG is very limited, however, high levels of *Aedes aegypti* resistance have been described. Here we present insecticide resistance monitoring data from across PNG generated between 2017 and 2022.

**Methods:** Mosquito larvae were collected in larval habitat surveys and through ovitraps. Mosquitoes were reared to adults and subjected to insecticide treated filter papers in WHO insecticide susceptibility bioassays. Subsets of *Aedes* mosquitoes were subjected to sequencing of the voltage-sensitive sodium channel (*Vssc*) region to identify resistance mutations.

**Results:** Overall, nearly 20,000 adult female mosquitoes from nine PNG provinces were used in the tests. We show that in general, Anopheline mosquitoes in PNG remain susceptible to pyrethroids but with worrying signs of reduced 24 h mortality in some areas. In addition, some *Anopheles* populations were indicated to be resistant against DDT. We show that *Ae. aegypti* in PNG are pyrethroid, DDT and likely bendiocarb resistant with a range of *Vssc* resistance mutations identified. We demonstrate that *Ae. albopictus* is DDT resistant and is likely developing pyrethroid resistance based on finding a low frequency of *Vssc* mutations.

**Conclusion:** This study represents the largest overview of insecticide resistance in PNG. While *Ae. aegypti* is highly pyrethroid resistant, the Anopheline and *Ae. albopictus* populations exhibit low levels of resistance in some areas. It is important to continue to monitor insecticide resistance in PNG and prepare for the widespread emergence of pyrethroid resistance in major disease vectors.

## Background

Insecticide resistance (IR) surveillance in mosquitoes is important for monitoring continued efficacy of insecticide-based vector control interventions and for guiding the selection and application of the most appropriate combinations of products and active ingredients [1, 2]. This includes the choice of products for long-lasting insecticidal nets (LLINs), indoor residual spraying, and other vector control tools. Ideally, a combination of products with active ingredients is used, against which no resistance in the population has been detected [3].

Papua New Guinea (PNG) is endemic for malaria and lymphatic filariasis transmitted by Anophelines [4, 5] and a wide range *of Aedes*-transmitted arboviruses including dengue, and chikungunya viruses [6, 7]. The majority of the *Anopheles* mosquito vector species in PNG belong to the *Anopheles (An*.*) punctulatus* complex including *An. farauti sensu stricto* (s.s), *An. koliensis* and *An. punctulatus* s.s. Minority species that have been demonstrated to transmit malaria include *An. longirostris* complex and *An. bancroftii* [8, 9]. The arbovirus vectors *Aedes (Ae*.*) aegypti* are found in larger, more densely populated centres whereas *Ae. albopictus* dominates in rural settings and smaller urban centres [10]. Locally important minority species include *Ae. scutellaris* [11, 12].

Over recent years we have reported IR status of malaria [13] and arbovirus [14] vectors in several locations in PNG, determined using the World Health Organization (WHO) insecticide susceptibility bioassays. Our previous data were limited to four provinces for *Anopheles* spp. and two provinces for *Aedes* spp. We reported full susceptibility of all tested Anophelines against pyrethroids (deltamethrin), carbamates (bendiocarb) and organophosphates (malathion), and potential resistance to organochlorides (dichlorodiphenyltrichloroethane (DDT))[13]. Furthermore, we reported severe pyrethroid resistance of *Ae. aegypti* and DDT resistance of *Ae. albopictus* found in Port Moresby and Madang. Genetic analysis indicated that all *Ae. aegypti* in PNG carried common target site resistance mutations conferring resistance to pyrethroids and DDT [14], but we were unable to pinpoint the genetic origin of DDT resistance in *Ae. albopictus* and the Anophelines.

Here we provide an update on IR monitoring in PNG that includes bioassay data from nine PNG provinces for *Anopheles* and *Aedes* mosquitoes conducted between 2017 and 2022. This update provides a comprehensive overview of the distribution and current insecticide resistance status in PNG disease vector populations. Our studies focus on the low altitude areas of PNG where vector-borne disease transmission is intense [6, 15].

## Materials and Methods

### Study site and mosquito sampling

The study was conducted across nine provinces of Papua New Guinea (Figure 1) with high burdens of vector-borne diseases transmission between 2017 and 2022. Specifically, the provinces included: National Capital District (NCD), Central Province, Milne Bay Province and Western Province in the southern region of mainland PNG; East New Britain and New Ireland in the New Guinea Islands region and East Sepik, Madang, Morobe, and West Sepik on the northern coast of mainland PNG. Most provinces were surveyed once, East Sepik Province was surveyed twice, and Madang Province was surveyed four times. Figure 1 shows a map of the surveyed provinces and the years the surveys were completed.

**Figure 1:**
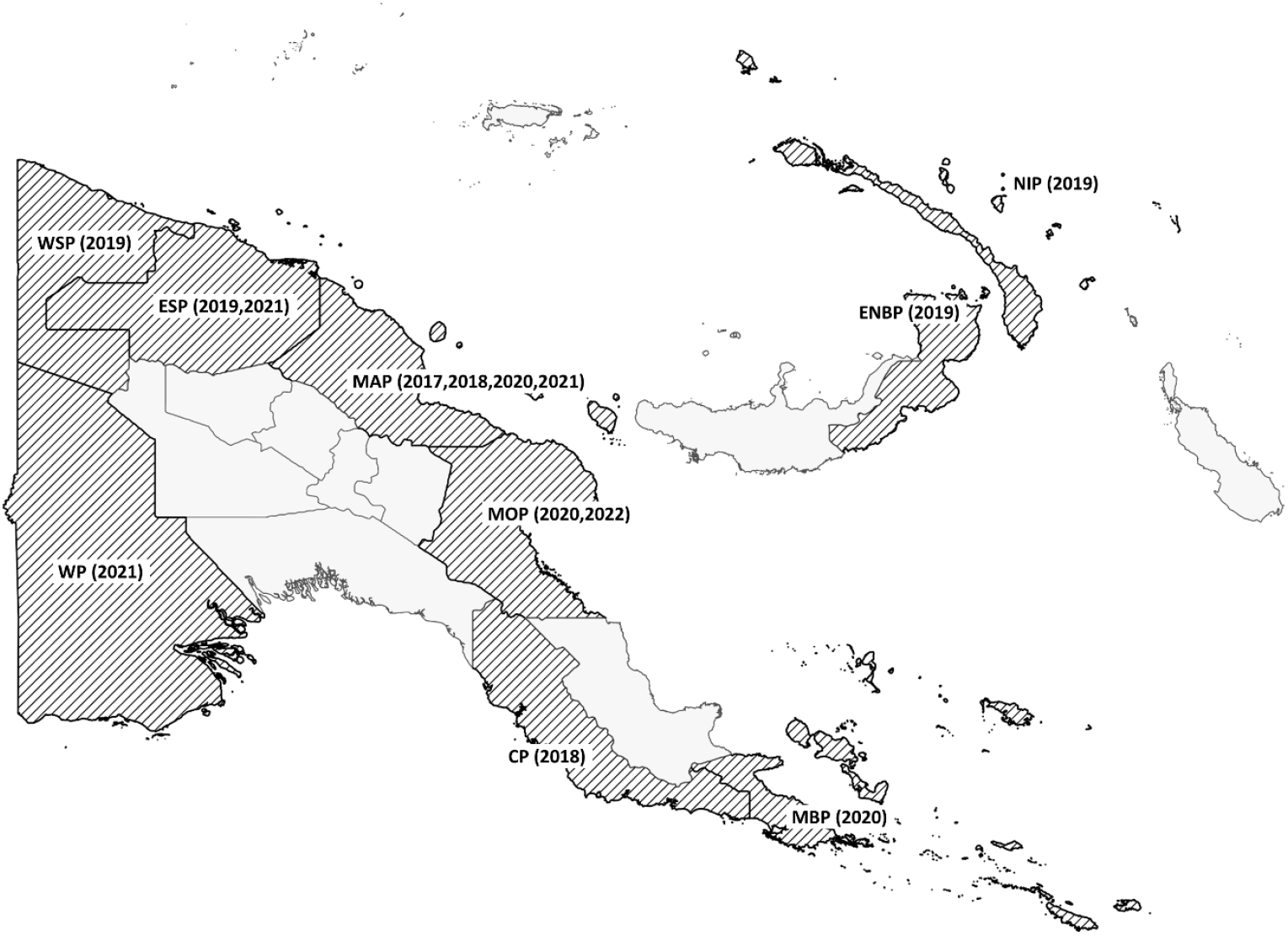
Map of Papua New Guinea and survey provinces. The provinces in which surveys were carried out are shaded and abbreviated as follows: National Capital District, Central Province (CP), Milne Bay Province (MBP), Western Province (WP), East New Britain (ENBP), New Ireland (NIP), East Sepik (ESP), Madang (MAP), Morobe (MOP), and West Sepik (WSP). The years when surveys were completed are shown in parentheses.

Provinces were surveyed for anopheline and aedine larvae at the same time. Anopheline larval habitats included drainage channels, sand-barred streams, transient puddles, forest swamps, pig-wallows, wells, riverine puddles. The collection of larvae in each province was limited by access via the road network and collections were conducted mainly along the sides of roads accessible by vehicle. On average, each survey included 14 days of larval collections. *Anopheles* larvae were collected, preferably as 3^rd^ or 4^th^ instars, from their habitats using plastic scoops, placed into 500 mL PETT bottles, labelled and brought back to a field insectary or to the PNG Institute of Medical Research (PNGIMR) insectary in Madang. Larval habitat characteristics of each sampled larval source were recorded, including Global Positioning System (GPS) locations. Data were entered into the electronic data capture system Epicollect5 (https://five.epicollect.net/; V4.2.0 Centre for Genomic Pathogen Surveillance, 2022) using mobile electronic tablets. For reference, tests were also conducted with a fully susceptible *An. farauti* colony maintained at PNGIMR [16].

*Aedes* eggs and larvae were collected in ovitraps [17, 18] and through larval habitat surveillance. A total of around 20 ovitraps per survey were set up across consenting households in an urban area (town or suburb) for five days. Each trap’s GPS location was logged electronically in EpiCollect5. Additionally, common aedine larval habitats were sampled and habitat characteristics recorded as described for anophelines.

### Mosquito Rearing

Temporary insectaries were set up for each survey except in Madang province where the PNGIMR has a permanent insectary. Mosquito larvae (both *Anopheles* spp. and *Aedes* spp.) were reared in shallow photo developing trays (sizes 16.5×11 cm or 24.5×18.5 cm or depending on the number of larvae collected per site, with larval density ranging from 200 to 500 per respective tray size.) The larvae were fed with ground Marine Master Tropical fish flakes (Marinepet Australia Pty Ltd). Pupae were collected daily into small plastic cups, and cups were placed into screened plastic containers (20 cm in diameter and 20 cm in height), or BugDorm-1 insect rearing cages (MegaView Science Co., Ltd, Taiwan, 1995-2022), with capacity to hold 300-500 adult mosquitoes at a time. Emerged adult mosquitoes were kept for 3-5 days in the cages before being used in WHO tube bioassays, with access to cotton balls soaked with 10% (w/v) sucrose solution placed on top of the cages and covered with damp cloth to keep ambient tropical temperature and humidity levels.

### WHO insecticide susceptibility bioassays

*Anopheles* and *Aedes* mosquitoes were tested against eight insecticides: pyrethroid type I insecticide, 0.75% permethrin; pyrethroid type II insecticides, 0.05% deltamethrin, 0.05% lambda-cyhalothrin and 0.05% alphacypermethrin; carbamate insecticide, 0.1% bendiocarb; organochloride insecticide, 4% DDT; and organophosphate insecticides, 5% malathion and, 0.25% pirimiphos-methyl. We also included the diagnostic concentrations of 0.03% deltamethrin and 0.8% malathion against *Aedes* populations in Morobe and NCD. These are concentrations unofficially recommended by WHO for testing *Aedes* mosquitoes [19]. The number of insecticides tested varied across surveys (subject to availability of mosquito larvae), with insecticides such as 0.05% deltamethrin and 4% DDT prioritized because of the current insecticide usage (deltamethrin LLINs) and historical usage of DDT in the country, across the nine provinces.

Bioassays were conducted using WHO standard procedures [2], ideally with 25 adult female anopheline mosquitoes per bioassay tube, and a full test having four replicates and two control tubes (n=150 adult female mosquitoes per test). However, if insufficient larvae were present to conduct a full test, tests were conducted with twenty to ninety mosquitoes (one to three replicates and 1 control per tests. Mosquitoes were exposed to insecticide-impregnated filter papers inside the replicate tubes for a total of 60 minutes, with knock down recorded at 5 minute intervals for 30 minutes, followed by 10 minute interval readings until 60 minutes. Mosquitoes were then transferred from each tube into separate holding tubes (controls included) and kept at an ambient tropical temperature and humidity for a 24-hour holding period (48 hours for 4% DDT), with access to 10% (w/v) sucrose solution. Mortality from each holding tube was recorded after 24 hours, with all survivors separated for easy identification during later laboratory analysis. Survivors and controls were knocked down at -20°C and morphologically identified using standard identification keys [20] along with the rest of the tested mosquitoes.

### Molecular Analysis

DNA was extracted from a sample of adult mosquitoes (*Ae. aegypti* n=55, *Ae. albopictus* n=170) using the Roche High Pure PCR template kit (Roche Molecular Systems, Inc., Pleasanton, CA, USA) according to the instructions of the manufacturer, but with two elution steps (first elution in 60 µL elution buffer and second elution in 120 µL). The second elution was diluted 1:10 with water and used in the molecular analysis of resistance mutations.

Custom TaqMan^®^ SNP Genotyping Assays (Life Technologies, California, USA) developed for each of the three target site mutations in the voltage sensitive sodium channel (*Vssc*) of *Aedes aegypti* (codons 989, 1016, 1534) were run on the Roche LightCycler® 480 and analysed using the endpoint genotyping method [21]. The *Vssc* amino acid positions are labelled as S989P, V1016G and F1534C according to the sequence of the most abundant splice variant of the house fly, *Musca domestica, Vssc* (GenBank accession nos. AAB47604 and AAB47605 [22]).

The *Vssc* codon 1534 of *Ae. albopictus* was screened for mutations using PCR primers aegSCF7/aegSCR7 and sequenced with aegSCR8. [23] 2 µL genomic DNA were amplified in a 25 µL PCR mix that included final concentrations of ThermoPol buffer Mg-free (1x) (New England Biolabs, Ipswich MA, USA), dNTPs (200 µM each), MgCl2 (1.5 mM), 0.5 µM each of forward and reverse primers, 0.625 units of Immolase™ Taq polymerase (Bioline, London, UK), and PCR-grade H2O. Thermocycling conditions followed those of Ahmad *et al*. [24]. PCR amplicons (740 bp) were sent to Macrogen Inc. (Seoul, South Korea) for Sanger sequencing on a 3730xl DNA analyser. Sequences were analysed, aligned, and trimmed (∼180 bp) using the program Geneious Prime® 2020.0.4 (Biomatters Ltd.).

### Data Analysis

Mosquito populations were classified as resistant or susceptible according to the WHO criteria [2]. Proportion of knocked down or dead mosquitoes was calculated and 24 h mortality was adjusted using control mortality and ‘Abbott’s formula’ where the control mortality was between 5% to 20%. Analysis of proportions (95% CI) and one-sample z-tests were conducted to determine if the observed 24 h mortality was indicative of resistance or susceptibility. Analyses were conducted for each species (where possible), insecticide and province.

## Results

### Species distribution of mosquitoes tested in the bioassays

A total of 11,210 anopheline mosquitoes and 8,294 aedine mosquitoes were used in the WHO tube bioassays across nine provinces in PNG between 2017 and 2022. The distribution of taxa identified based on morphology is shown in Figure 2. Bioassays were also conducted with a pyrethroid susceptible *An. farauti* colony at PNGIMR Madang (n = 1,762).

**Figure 2:**
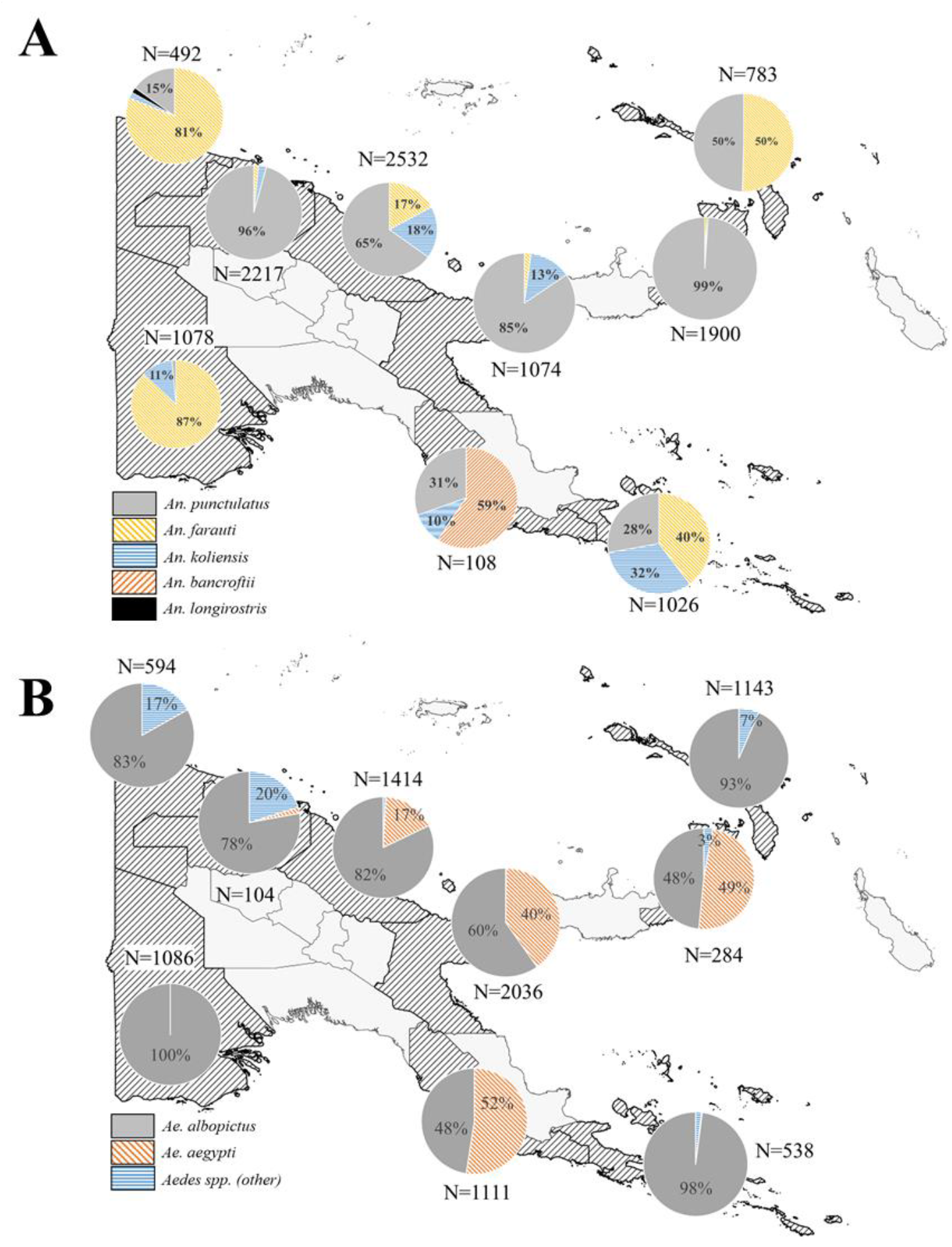
Mosquito species distribution used in WHO tube bioassays. Panel A: Anopheline species; Panel B: *Aedes* species. Percentages below 2% are not explicitly presented in the pie charts. The total number of mosquitoes exposed in the bioassays per province is also shown (N).

### Overall insecticide resistance profile of PNG vector species

Table 1 shows the overall insecticide susceptibility profiles of *An. farauti* colony mosquitoes, *An. punctulatus* s.l, *Ae. albopictus, Ae. aegypti*, and minority species (combined) as determined in the present study.

**Table 1:** Summary of phenotypic resistance bioassay results of the major malaria and arbovirus vector species in PNG. Results are shown as 24 h mortality in percent with 95% confidence intervals given (95% CI) along with the total number of mosquitoes exposed [N]. Cells indicating a resistant phenotype are highlighted in red and cells indicating a suspected resistant phenotype are highlighted in yellow. * Insecticide susceptible *An. farauti s*.*s* colony maintained at PNGIMR. NA indicates that sample numbers were too low or not tested.

| *Insecticide* | *An. farauti* s.s*<br>% (95% CI) [N] | *An. punctulatus* s.l<br>% (95% CI) [N] | *An. spp.*<br>% (95% CI) [N] | *Ae. albopictus*<br>% (95% CI) [N] | *Ae. aegypti*<br>% (95% CI) [N] | *Ae. spp.*<br>% (95% CI) [N] |
| --- | --- | --- | --- | --- | --- | --- |
| 0.03%<br>deltamethrin | NA | NA | NA | **97.0**<br>(90-99) [87] | **20.2**<br>(13.0-29.0) [99] | NA |
| 0.05%<br>deltamethrin | **100**<br>(99.2-100) [487] | **99.5**<br>(99.2-99.8) [2241] | **100**<br>(92.1-100) [45] | **98.7**<br>(97.8-99.3) [1059] | **31.8**<br>(26.9-37.0) [346] | **100**<br>(92.6-100) [48] |
| 0.05%<br>lambdacyhalothrin | **100**<br>(98.0-100) [186] | **99.2**<br>(98.5-99.8) [757] | NA | **99.7**<br>(98.6-100) [387] | **32.0**<br>(19.5-46.7) [50] | **100**<br>(76.8-100) [14] |
| 0.05%<br>alphacypermethrin | **100**<br>(96.4-100) [101] | **100**<br>(97.9-100) [172] | NA | **100**<br>(94.2-100) [62] | **64.3%**<br>(48.0-78.5)[42] | NA |
| 0.75%<br>permethrin | **100**<br>(96.3-100) [99] | **100**<br>(96.6-100) [106] | NA | **100**<br>(95.6-100) [88] | **10.7**<br>(2.3-28.2) [28] | NA |
| 4%<br>DDT | **100**<br>(96.2-100) [94] | **95.9**<br>(94.9-96.8) [1812] | NA | **78.5**<br>(75.4-81.4) [762] | **18.9**<br>(12.5-26.8) [127] | **89.5**<br>(66.9-98.7) [19] |
| 0.8%<br>malathion | NA | NA | NA | **43.0**<br>(36-50) [193] | **62.8**<br>(55.0-70.0) [164] |  |
| 5%<br>malathion | **100**<br>(96.3-100) [97] | **100**<br>(99.7-100) [1051] |  | **97.2**<br>(95.5-98.4) [567] | **99.5**<br>(97.0-100.0) [203] | **100**<br>(93.0-100) [51] |
| 0.25%<br>pirimiphos-methyl | **100**<br>(96.8-100) [114] | **100**<br>(98.7-100) [287] | NA | **99.2**<br>(95.8-100) [129] | **100**<br>(91.0-100) [39] | NA |
| 0.1%<br>bendiocarb | **100**<br>(96.2-100) [96] | **99.9**<br>(99.5-100) [1177] | NA | **98.5**<br>(97.5-99.2) [922] | **94.0**<br>(88.0-97.0) [133] | **100**<br>(92.1-100) [45] |

*An. punctulatus* s.l populations (Table 1) showed susceptibility to all tested insecticides at discriminating concentrations except for 4% DDT, for which resistance was indicated, with 95.8% (95% CI: 94.8-96.7%) 24 h mortality. *An*. spp. (mainly *An. bancroftii* and *An. longirostris*) numbers were too low to draw any conclusions for most insecticides except for 0.05% deltamethrin, where susceptibility was indicated.

*Ae. aegypti* populations present in urban centres across PNG exhibit high levels of pyrethroid and DDT resistance as indicated by an average 24 h mortality ranging from 20%-32% for discriminating concentrations of deltamethrin and lambda-cyhalothrin and 15% (95% CI: 9-23%) for DDT (Table 1). This was based on the 0.05% deltamethrin and 0.03% deltamethrin (WHO recommended) discriminating concentrations for *Ae. aegypti*.

*Ae aegypti* also showed resistance to malathion at the WHO recommended discriminatory concentration of 0.8% but showed high mortality at the higher 5% concentration recommended for Anophelines. This concentration is approximately six times higher than the interim recommended *Aedes* discriminatory concentration. According to the WHO intensity bioassay criteria [2], the species when combined across PNG shows a low level of resistance intensity. Resistance against bendiocarb was also indicated with 94% mortality after 24 hours (95% CI: 88-97%).

*Ae. albopictus* showed resistance to DDT (77%, 95% CI: 74-80%) and to the 0.8% malathion (43%, 95% CI: 36-50%) discriminating concentration, and possible resistance to 0.03% deltamethrin (97%, 95% CI: 90-99%), and 5% malathion (97%, 95% CI: 97-100%). *Ae. albopictus* showed susceptibility to 0.05% deltamethrin (99%, 95% CI: 98-100%), lambdacyhalothrin (100%), permethrin (100%) and 0.1% bendiocarb (99%, 95% CI: 98-100%).

Other *Aedes* species (which are mainly *Ae. scutellaris* from the northern PNG) were present in numbers too low to draw definitive conclusions. However, results suggest phenotypic resistance against DDT with 24 h mortality of 80% (95% CI: 56.3-94.3%).

### Spatial and species-specific results of Anopheles punctulatus s.l morphospecies

Species-specific 24 h mortality for the three major *Anopheles* morphospecies *An. farauti, An. punctulatus* s.s. and *An. koliensis* in each province is shown in Figure 3. When data were stratified by morphospecies and province, sample size usually became quite small, limiting the confidence of inferences made from the analyses.

**Figure 3:**
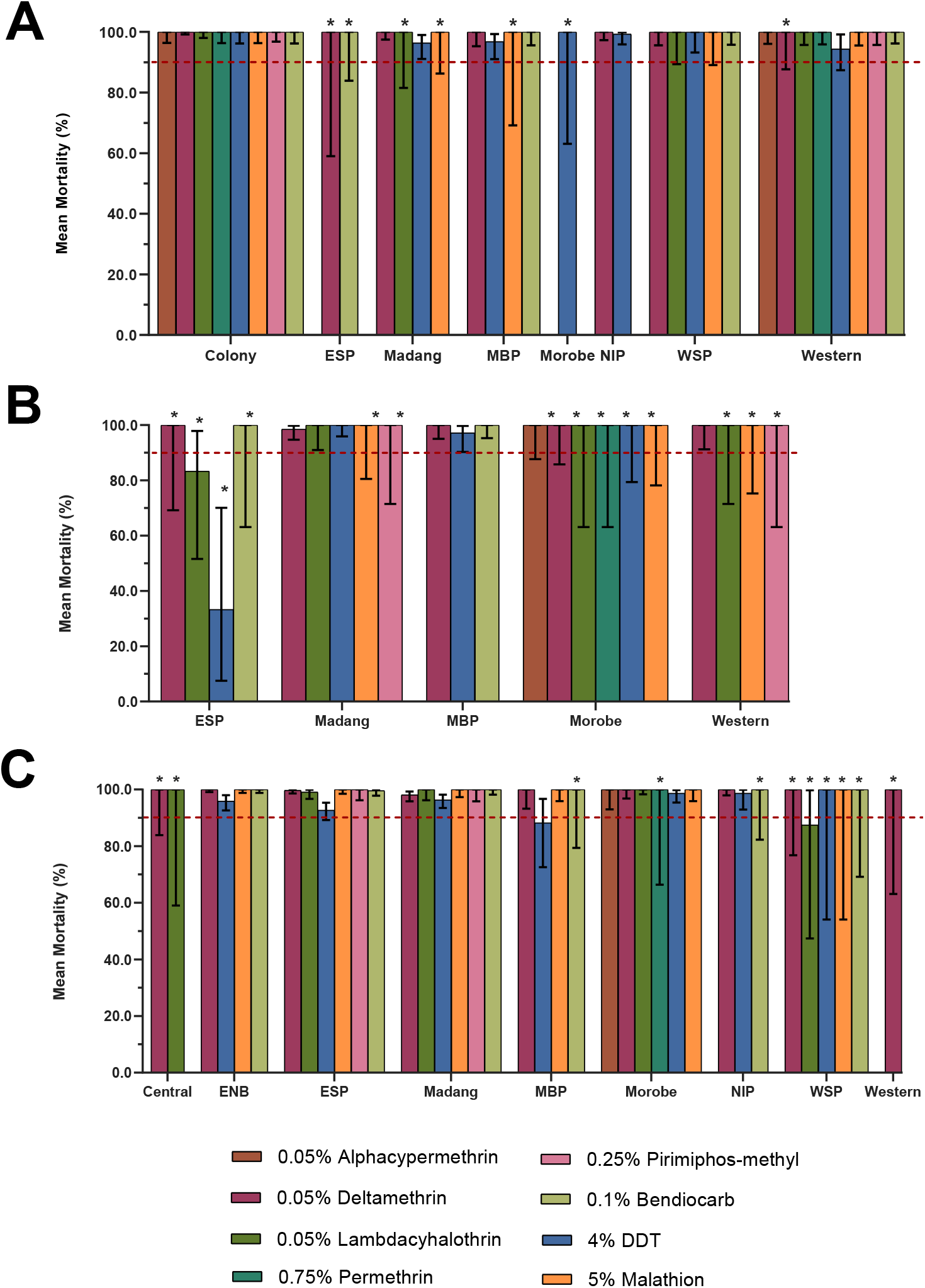
Mean mortality rates of the *Anopheles punctulatus* complex (*An. punctulatus* s.l) for nine selected provinces of PNG, against eight different insecticides. Panel A: *Anopheles farauti*, Panel B: *Anopheles koliensis*; Panel C: *Anopheles punctulatus* s.s. Provinces surveyed indicated on the x-axis, abbreviations are; East New Britain (ENB), East Sepik (ESP), Milne Bay (MBP), New Ireland (NIP) and West Sepik (WSP). 95% CIs indicated by error bars. The WHO resistance threshold line for discriminatory doses shown at 90% mean mortality. Sample numbers ≤ 5 not included and > 5 ≤ 30 indicated with asterisks. Controls not included.

Data indicate susceptibility against all tested insecticides in all morphospecies and provinces, including deltamethrin which has been only insecticide used in long-lasting insecticidal nets (LLINs) in PNG between 2005 and 2019 [25, 26]. Results from the *An. koliensis* population in East Sepik province indicate higher than average levels of DDT and lambda-cyhalothrin resistance, with 24 h mortality of 33.3% (95% CI: 7.49-70.1%) and 83.3% (95% CI: 51.6 - 97.7%) respectively. Results from the *An. punctulatus* population also indicate higher than average levels of DDT resistance in Milne Bay province, with a 24 h mortality of 88.2% (95% CI: 72.6-96.7%) and lambda-cyhalothrin resistance in West Sepik province, 24 h mortality of 87.5% (95% CI: 47.4-99.7%).

DDT results also indicated possible resistance in the *An. farauti* population in Madang, Milne Bay and Western provinces with the 24 h mortality rates ranging between 94%-97%, and in the *An. punctulatus* population of East New Britain, East Sepik and Madang provinces, with 24 h mortality rates ranging between 92%-97% (Figure 3).

### Spatial and species-specific results for Aedes aegypti and Ae. albopictus

Species-specific 24 h mortality for the two major *Aedes* species *Ae. aegypti* and *Ae. albopictus*, in each province is shown in Figure 4.

**Figure 4:**
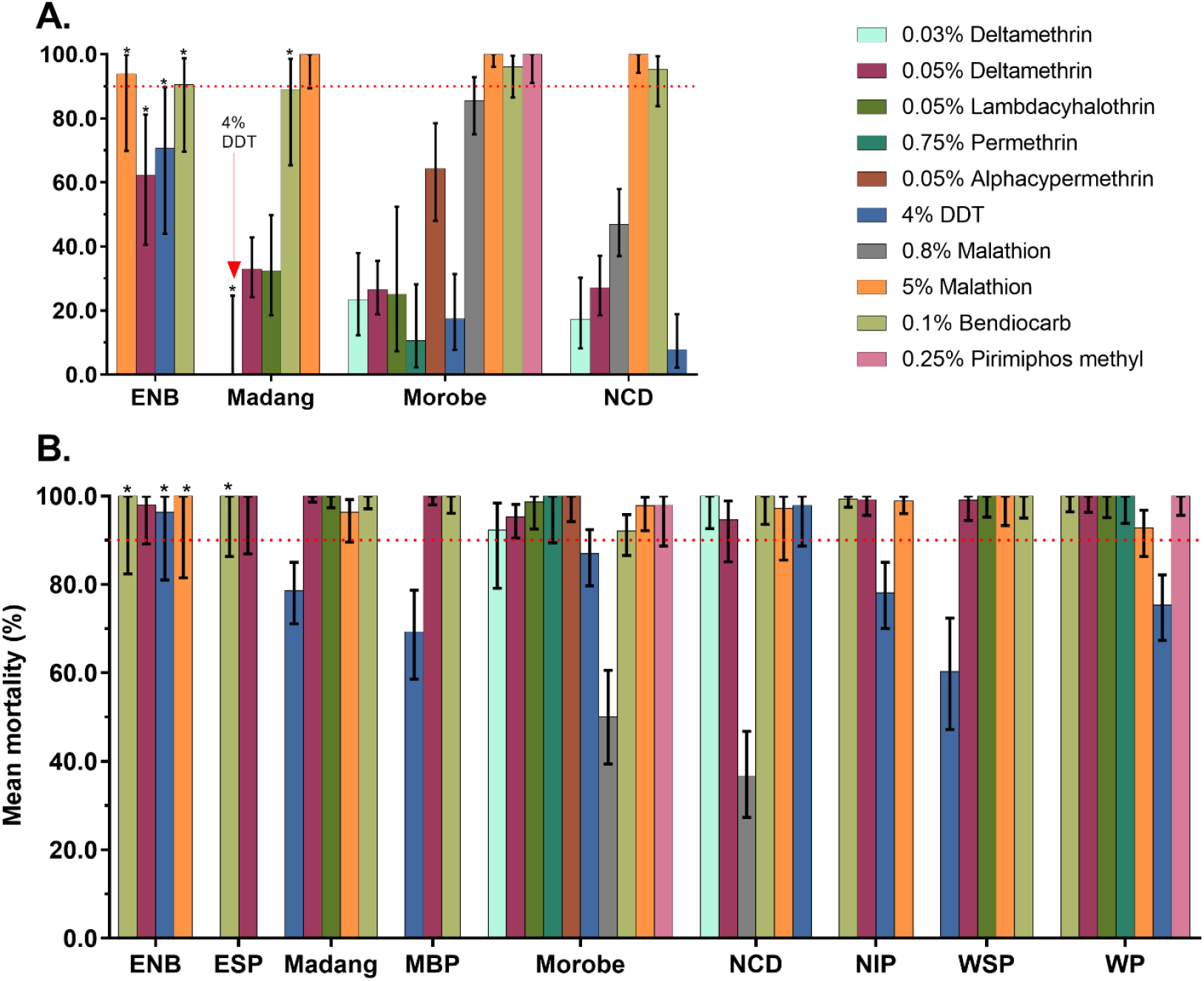
Mean mortality rates of populations of *Ae. aegypti* (panel A) and *Ae. albopictus* (panel B) across different provinces in PNG against different insecticides. Error bars are 95% confidence intervals. *Low sample size (<30 mosquitoes), however, mortality rates are displayed to show the landscape of insecticide susceptibility across PNG.

**Figure 5:**
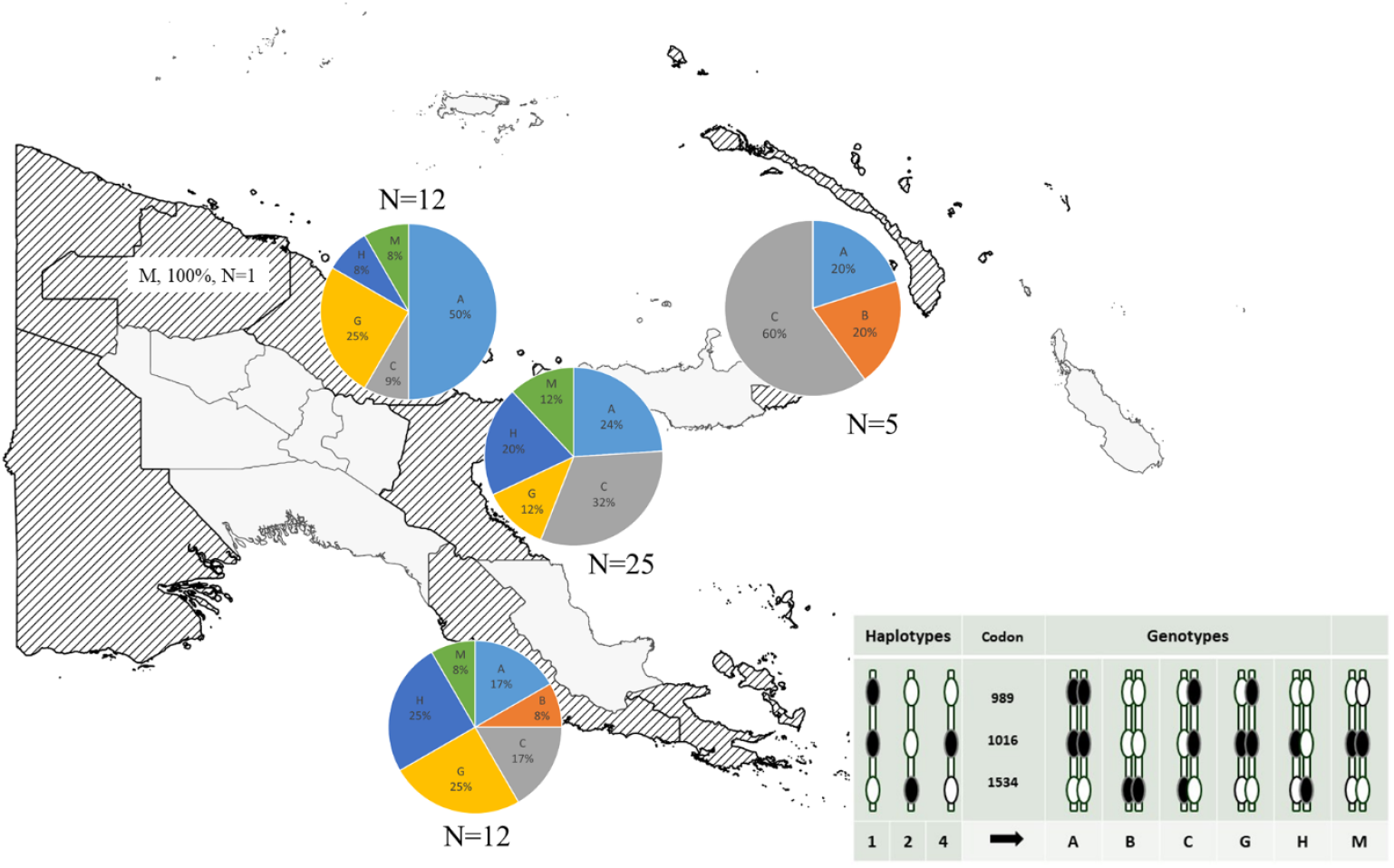
Distribution of *kdr* mutations in n=55 *Aedes aegypti* samples from 5 provinces in PNG. The legend shows the haplotypes and resulting genotypes that were identified. Each genotype was assigned a letter. The size of the pie charts is arbitrary and not reflecting sample size. East Sepik Province only had 1 sample, therefore no pie chart is presented.

*Aedes aegypti* were found in four population centres, namely Port Moresby (NCD), Lae (Morobe), Madang (Madang Province), and in Kokopo and Rabaul (East New Britain Province). Variable resistance levels were observed to deltamethrin, lambdacyhalothrin, DDT, malathion and bendiocarb (Figure 4). Specifically, high deltamethrin resistance was observed at WHO-recommended discriminatory concentration for *Aedes aegypti* of 0.03% ranging from 17% to 0.23% 24 h mortality in Morobe and NCD. At the higher discriminatory concentration recommended for *Anopheles* mosquitoes (0.05 %) tested across all four *Ae. aegypti* populations, we observed between 17%-63% 24 h mortality. Observed 24 h mortalities for 0.03% and 0.05% deltamethrin discriminatory concentrations were not significantly different in NCD and Morobe. Mortalities against the 0.05% deltamethrin discriminatory concentration were lower in Madang, NCD and Morobe as compared to East New Britain province. Lambdacyhothrin resistance was tested only in ESP and indicated with 32% (95% CI: 0.18-0.50) 24 h mortality. High levels of DDT resistance with 24 h mortality ranging from 0% to 71% were detected in across all four provinces. There was no difference between 24 h and 48 h mortality rates for this AI. Similar to deltamethrin, we observed significantly lower 24 h mortality rates against 4% DDT in East New Britain as compared to the other three provinces where *Ae. aegypti* was found. Malathion resistance to the 0.8% malathion discriminatory concentration recommended by WHO for *Aedes spp*. was detected in Morobe with 86% dead (95% CI: 75%-93%) and NCD with 47% dead (95% CI: 37%-58%). At the higher concentration of 5% malathion, Morobe, NCD and Madang were fully susceptible, but possible resistance was detected in East New Britain with 24 h mortality of 94% (95% CI: 70%-100%). In Morobe and NCD, we show that the resistance intensity level was low. Possible resistance to bendiocarb with 24 h mortality ranging from 89-95% was detected in Morobe, NCD, ENB and Madang.

*Ae. albopictus* was found in all nine provinces surveyed. Populations across the nine provinces showed mainly susceptibility or possible low grade resistance against the panel of insecticides tested except for DDT, for which higher levels of resistance were observed (Table 1). DDT resistance was indicated by 24 h mortality rates between 60% and 79% in West Sepik, Milne Bay, Western, Morobe, New Ireland and Madang. Possible resistance was observed in East New Britain with 96% (95% CI: 81%-100%) mortality and in NCD with 98% (95% CI: 89%-100%) mortality. The deltamethrin discriminating concentration of 0.03% tested in Morobe and NCD showed *Ae. albopictus* resistance in the former at a low grade with 92% (95% CI:79-98%) 24 h mortality, but full susceptibility in NCD. All *Ae. albopictus* in the seven provinces outside of Morobe and NCD were susceptible with percentages dead ranging from 98-100%. The species was fully susceptible to the other pyrethroid insecticides, lambdacyhalothrin, in Madang, WSP and Western (100% 24 h mortality). Western population was also fully susceptible to permethrin. Malathion resistance was observed at the much lower discriminating concentration of 0.8% recommended by WHO with in Morobe 50% (95% CI:39%-61%) and in NCD with 37% (95% CI:27-47%) 24 h mortality. At the higher concentration of 5%, we observed 24 h mortality of 97% in both populations, which may indicate a moderate to high resistance intensity to malathion. There also was possible malathion resistance detected using the 5% discriminatory concentration in Madang with 96% (95% CI: 090-99%) mortality and in Western with 93% (95% CI: 86-97%). *Ae. albopictus* in New Ireland and WSP were fully susceptible to 5% malathion, with 24 h mortalities between 99% and 100%. Bendiocarb susceptibility was indicated with 99-100% 24 h mortality in all provinces except in Morobe province where a low-grade resistance was indicated with 91% (95% CI: 0.80-0.97) 24 h mortality. Pirimiphos-methyl was only tested in Western Province and resulted in 100% 24 h mortality.

### Molecular Analyses in *Aedes aegypti* and *Ae. albopictus*

Mutations in the *Vssc* gene in *Ae. aegypti* were found to be common at codons 1016, 1534 and 989 in the PNG sample. Six composite *Vssc* genotypes arising from four haplotypes were identified in the n=55 samples of *Ae. aegypti* from five PNG provinces (Table 2). Three of the genotypes are known to confer pyrethroid resistance and it is likely that the remaining three would also confer some level of resistance though they have not all been tested specifically. No wild-type genotype or haplotype was identified in the sample.

**Table 2:** Sequencing results for the region including codon 1534 of the *Vssc* in *Aedes albopictus*. Most samples were wildtype but low frequencies of resistance mutations were identified in Milne Bay, Morobe and Western Provinces.

| Province | 1534 |  | n | freq in Province |
| --- | --- | --- | --- | --- |
| East New Britain | TT | WILDTYPE | 26 | 1.00 |
| East Sepik | TT | WILDTYPE | 44 | 1.00 |
| Milne Bay | TT | WILDTYPE | 9 | 0.82 |
| Milne Bay | TG | F1534C | 1 | 0.09 |
| Milne Bay | GG | 1534C | 1 | 0.09 |
| Morobe | TT | WILDTYPE | 20 | 0.91 |
| Morobe | TG | F1534C | 2 | 0.09 |
| New Ireland | TT | WILDTYPE | 12 | 1.00 |
| West Sepik | TT | WILDTYPE | 10 | 1.00 |
| Western | TT | WILDTYPE | 41 | 0.98 |
| Western | TG | F1534C | 1 | 0.02 |

The most frequent genotype (frequency = 0.27) in the sample was a homozygous mutation at codon 1016 and 989 with the wildtype homozygote at codon 1534. Another common genotype found was the triple heterozygote at V1016G, F1534C and S989P (genotype frequency = 0.26). A new genotype (1016G in the homozygous state and not linked to 989P - now known as genotype M) for PNG was found at a frequency of 11% in the sample. The distribution of *Vssc* genotypes across the sampled provinces of PNG revealed resistance mutations to be widespread.

**Table 1:** Six composite genotypes identified in *Aedes aegypti* in PNG.

| Codon |  |  | n | frequency |
| --- | --- | --- | --- | --- |
| 1016 | 1534 | 989 |  |  |
| GG | TT | CC | 15 | 0.273 |
| TT | GG | TT | 2 | 0.036 |
| TG | TG | TC | 14 | 0.255 |
| GG | TT | TC | 9 | 0.164 |
| TG | TG | TT | 9 | 0.164 |
| GG | TT | TT | 6 | 0.109 |

Genotyping by sequencing the region including codon 1534 of the *Vssc* in *Aedes albopictus* showed that most mosquitoes in the sample were wildtype for the resistance mutation. However, one homozygote and four heterozygotes for this mutation were found in a sample of 166 individuals. The mosquitoes carrying this mutation were collected in Milne Bay and Morobe Provinces (Table 2).

## Discussion and Conclusion

The present study provides the largest currently available dataset on insecticide resistance status of malaria and arbovirus vectors in PNG. Using standard WHO tube bioassays, we characterized phenotypic resistance of *Anopheles* and *Aedes* species against important insecticides used in public health programs in nine provinces experiencing high vector-borne disease burden.

The three major malaria vector species, *An. farauti, An. punctulatus s*.*s*. and *An. koliensis*, have been previously profiled using the same methods as susceptible to deltamethrin, lambda-cyhalothrin and DDT in Madang, East Sepik, Milne Bay and East New Britain provinces [13].

Susceptibility of *An. punctulatus s*.*l* to the AIs used in malaria control in PNG, in particular deltamethrin, which has been the single AI used in LLINs in PNG for over 15 years, indicates a surprising longevity of these tools in PNG.

We show here that the *An. punctulatus* in ESP and ENB are showing signs of emerging phenotypic resistance to DDT and lambda-cyhalothrin. If cross resistance to deltamethrin and or alphacypermethrin arises, this may compromise the effectiveness of purely pyrethroid-based malaria control strategies in PNG in the future.

Our data also indicate DDT resistance in *An. farauti* and *An. koliensis*, however this could not be conclusively shown due to the low numbers of the species tested in ESP. *An. koliensis* was reportedly resistant to DDT in neighboring Irian Jaya, Indonesia [27]. It is imperative that future insecticide resistance surveillance confirm the resistance status of these important malaria vector species.

Whether the observed signs of DDT resistance in Anopheline populations in some areas are a remnant of historic DDT usage in PNG [28] or emerging cross-resistance with pyrethroids [29] is yet to be elucidated. The second hypothesis is supported by our data showing resistance against lambda-cyhalothrin in some provinces. Furthermore, the cross-resistance profile (DDT-lambdacyhalothrin) in the South American malaria vector, *An. darling*, conferred through a metabolic detoxification pathway [30], could potentially be driving this phenomenon in our local vector populations in addition to target site mutations.

There is an urgent need to confirm the underlying resistance mechanisms using molecular analysis tools such as *Vssc* genotyping and population genomic tools such as Whole Genome Sequencing and investigation of gene copy number variation[31]. There is also a role for expanding bioassay tools to include use of synergists to identify components of metabolic resistance. Widespread emergence of pyrethroid resistance in PNG would compromise current public health vector control strategies.

*Ae. aegypti* and *Ae. albopictus* are the two common arboviral vectors in PNG. Previous profiling of these species in Madang and Port Moresby (NCD) reported pyrethroid resistance in *Ae. aegypti* in Port Moresby (NCD) and Madang. DDT resistance was detected in *Ae. albopictus* in Madang. While both species in the two areas were susceptible to malathion, *Ae. aegypti* indicated resistance to DDT and bendiocarb (not confirmed due to low sample size) and *Ae. albopictus* was susceptible to lambdacyhalothrin [14].

The emergence of resistance to deltamethrin, lambdacyhalothrin, DDT and malathion in *Ae. aegypti* and *Ae. albopictus*, confirmed in several populations, highlights a critical need to understand the landscape of chemical use for pest control (private sector, public health, agricultural or industrial) in PNG. DDT was last used almost 50 years ago in a national malaria control program [32], however it has been reported to be used illegally within the country for either public health or agriculture [33]. Products containing a number of AIs including malathion and lambdacyhalothrin are frequently used in PNG [33] suggesting that selective pressure from these AIs could be contributing to the high resistance in *Ae. aegypti* and the emergence in *Ae. albopictus*.

The capital cities of Port Moresby (NCD) and Lae (Morobe) are the most urbanized and densely populated areas in PNG. Both *Aedes* species thrive in these environments at high densities similar to elsewhere in Asia [34], Latin America [35], and Africa [10], posing a high risk of arboviral transmission, in particular dengue fever, chikungunya and Zika within these cities. Widespread resistance in *Ae. aegypti* and *Ae. albopictus* to multiple insecticides in these provinces (Morobe and NCD) highlights a critical need for a harmonized vector control plan that takes into account these resistance profiles. Furthermore, building capacity for routine mosquito surveillance in PNG and strengthening partnerships across PNG research institutions, government departments and health laboratories is critical to reduce these important mosquito vector populations and thus reduce disease transmission [36].

The intense pyrethroid resistance within the *Ae. aegypti* populations in PNG is conferred, at least in part, by three or more different *kdr* genotypes. The most frequent genotype, a homozygous mutation at codon 1016 and 989 with the wildtype homozygote at codon 1534, is a common genotype also found in Bali and other locations throughout Southeast Asia and the Pacific, and confers a high level of resistance to Type I and II pyrethroids [21, 37]. The heterozygote at V1016G, F1534C and S989P is found commonly in *Ae. aegypti* in other countries in the region and confers a low to moderate level of resistance to Type I and II pyrethroids [38]. The homozygote 1534C occurring alone is rare in the PNG sample, but confers a low level resistance to Type I pyrethroids [38]. A new genotype for PNG (1016G in the homozygous state and not linked to 989P - now known as genotype M) has not been found in recent studies of *Ae. aegypti* in the Indo-Pacific region [21], but does exist in Taiwan and its association with resistance has not been ascertained [39]. The two remaining genotypes identified in this study contain the haplotype that makes up genotype M and their association with pyrethroid resistance is yet to be tested. The role of metabolic resistance in the PNG mosquito populations has not been investigated. For both *Aedes* species studied in PNG, it will be important to determine whether the *Vssc* resistance mutations have arisen due to local selection or have arrived with invasive mosquitoes as has been shown in other parts of the Indo-Pacific region [21]. Whatever their origin, the continued presence of the *Vssc* mutations implies that they are being maintained due to selection with pyrethroid insecticides in PNG as there are known fitness costs associated at least with the 1016G/989P mutations [40, 41].

## Conclusion

We show that *An. punctulatus*, an important malaria vector species is resistant to DDT, with a possible cross resistance to the pyrethroid insecticide, lambdacyhalothrin. Further monitoring of the insecticide resistance status of other malaria vector species needs to be maintained. Vector control of arbovirus vector species such as *Ae. aegypti* in PNG needs to be streamlined and regulated under one regulatory body especially at provincial level. The existing toolbox for controlling these vectors is compromised due to the high severity of pyrethroid, organochloride and organophosphate resistance. Monitoring of all the important vector species at both phenotypic and molecular levels is crucial to identify and develop effective strategies in a timely manner to mitigate the spread of resistance across PNG.

## Data Availability

All data produced in the present study are available upon reasonable request to the authors

## Acknowledgements

We would like to thank the PNG communities where mosquito samples were collected for their support. We acknowledge the support from Provincial Health Authorities in all surveyed provinces. We thank the PNG National Malaria Control Program (NMCP), Rotarians Against Malaria PNG, and the PNGIMR Project Management Unit for the Global Fund Grant for management support, especially Tim Freeman, Munir Ahmed, Michelle Auryan, Sharon Jamea and Serah Kurumop. We thank the PNG National Agriculture and Quarantine Inspection Authority, especially David Tenakanai and Orlando Mercado for enabling operational support in several provinces. We acknowledge the Lihir Malaria Elimination Project team and the vector control unit International ISOS (Newcrest Lihir) for operational support, especially Pere Millat, Barbara Baro and Tony Tandrapah.

We thank current and former staff of the Entomology section at the PNGIMR Vector Borne Diseases Unit in Madang and the PNGIMR Maprik branch for contributions to logistical organization, sample collection and WHO tube bioassays, especially Michael Kurumop, Lincoln Timinao, Rebecca Vinit, Elias Omera, Peter Kaman, Benson Kiniboro and Kenny Rupa. We thank Rachael Farquhar (Burnet Institute) and Annie Dori (PNG NDoH) for STRIVE PNG partnership coordination and logistics support. We thank Tanya Russell (JCU) for support and technical guidance, and with drafting of standard operating procedures. We thank the STRIVE PNG Vector Working Group and the Technical Working Group of the NMCP, especially Tom Burkot, Nigel Beebe, Trevor Kelebi, Enoch Waipeli, Lucy Dally, Melinda Susapu and John Deli for helpful discussions and guidance.

## Author Contribution

Conceived study: SK, LM, ML, LJR, AH; Sample Collection and Experiments: MK, SL, NEH, SD, JG, NV, MS, AD, LK, SG, CP, SK; Data Analysis: MK, SL, NEH, SK; Wrote first draft: MK, SL, SK; Reviewed manuscript: MK, SL, NEH, AH, LM, LJR, ML, SK.

